# Prediction of post-PCV13 pneumococcal evolution using invasive disease data enhanced by inverse-invasiveness weighting

**DOI:** 10.1101/2023.12.10.23299786

**Authors:** Xueting Qiu, Lesley McGee, Laura L Hammitt, Lindsay R Grant, Katherine L O’Brien, William P Hanage, Marc Lipsitch

## Abstract

**Background:** After introduction of pneumococcal conjugate vaccines (PCVs), serotype replacement occurred in the population of *Streptococcus pneumoniae.* Predicting which pneumococcal clones and serotypes will become more common in carriage after vaccination can enhance vaccine design and public health interventions, while also improving our understanding of pneumococcal evolution. We sought to use invasive disease data to assess how well negative frequency-dependent selection (NFDS) models could explain pneumococcal carriage population evolution in the post-PCV13 epoch by weighting invasive data to approximate strain proportions in the carriage population.

**Methods:** Invasive pneumococcal isolates were collected and sequenced during 1998–2018 by the Active Bacterial Core surveillance (ABCs) from the Centers for Disease Control and Prevention (CDC). To predict the post-PCV13 population dynamics in the carriage population using a NFDS model, all genomic data were processed under a bioinformatic pipeline of assembly, annotation, and pangenome analysis to define genetically similar sequence clusters (i.e., strains) and a set of accessory genes present in 5% to 95% of the isolates. The NFDS model predicted the strain proportion by calculating the post-vaccine strain composition in the weighted invasive disease population that would best match pre-vaccine accessory gene frequencies. To overcome the biases of invasive disease data, serotype-specific inverse-invasiveness weights were defined as the ratio of the proportion of the serotype in the carriage data to the proportion in the invasive data, using data from 1998–2001 in the United States, before conjugate vaccine introduction. The weights were applied to adjust both the observed strain proportion and the accessory gene frequencies.

**Results:** Inverse-invasiveness weighting increased the correlation of accessory gene frequencies between invasive and carriage data with reduced residuals in linear or logit scale for pre-vaccine, post-PCV7, and post-PCV13. Similarly, weighting increased the correlation of accessory gene frequencies between different time periods in the invasive data. By weighting the invasive data, we were able to use the NFDS model to predict strain proportions in the carriage population in the post-PCV13 epoch, with the adjusted R-squared between predicted and observed strain proportions increasing from 0.176 to 0.544 after weighting.

**Conclusions:** The weighting system adjusted the invasive disease surveillance data to better represent the carriage population of *S. pneumoniae*. The NFDS mechanism predicted the strain proportions in the projected carriage population as estimated from the weighted invasive disease frequencies in the post-PCV13 epoch. Our methods enrich the value of genomic sequences from invasive disease surveillance, which is readily available, easy to collect, and of direct interest to public health.

**IMPORTANCE:** *Streptococcus pneumoniae*, a common colonizer in the human nasopharynx, can cause invasive diseases including pneumonia, bacteremia, and meningitis mostly in children under 5 years or older adults. The PCV7 was introduced in 2000 in the United States within the pediatric population to prevent disease and reduce deaths, followed by PCV13 in 2010, PCV15 in 2022, and PCV20 in 2023. After the removal of vaccine serotypes, the prevalence of carriage remained stable as the vacated pediatric ecological niche was filled with certain non-vaccine serotypes. Predicting which pneumococcal clones, and which serotypes, will be most successful in colonization after vaccination can enhance vaccine design and public health interventions, while also improving our understanding of pneumococcal evolution. While carriage data, which are collected from the pneumococcal population that is competing to colonize and transmit, are most directly relevant to evolutionary studies, invasive disease data are often more plentiful. Previously, evolutionary models based on negative frequency-dependent selection (NFDS) on the accessory genome were shown to predict which non-vaccine strains and serotypes were most successful in colonization following the introduction of PCV7. Here, we show that an inverse-invasiveness weighting system applied to invasive disease surveillance data allows the NFDS model to predict strain proportions in the projected carriage population in the post-PCV13/pre-PCV15 and -PCV20 epoch. The significance of our research lies in using a sample of invasive disease surveillance data to extend the use of NFDS as an evolutionary mechanism to predict post-PCV13 population dynamics. This has shown that we can correct for biased sampling that arises from differences in virulence and can enrich the value of genomic data from disease surveillance and advances our understanding of how NFDS impacts carriage population dynamics after both PCV7 and PCV13 vaccination.

## Introduction

Understanding pathogen population response to vaccination is essential for forecasting disease burden and improving vaccine composition, especially for pathogens that evolve quickly and/or are characterized by cocirculating antigenically diverse strains. *Streptococcus pneumoniae* (the pneumococcus), a common nasopharyngeal colonizer in children, is such a pathogen which has been categorized into more than 100 serotypes. Invasive pneumococcal disease (IPD), including pneumococcal pneumonia, bacteremia, and meningitis has historically been a leading source of pediatric morbidity and mortality. Since pneumococcal conjugate vaccine (PCV) was introduced in children in 2000, PCVs have prevented many cases of IPD and non-invasive disease in both children and indirectly adults, and produced significant public health benefits globally (1, 2). Because PCVs target only a small number of the more than 100 pneumococcal serotypes, the non-vaccine serotypes (NVTs) have increased in prevalence and partially eroded the benefits of vaccination. The prevalence of NVTs nasopharyngeal carriage has increased substantially among children after the introduction of 7-valent PCV (PCV7) with little or no net change observed in the carriage prevalence of the bacteria (3). The increase in NVTs’ absolute abundance alongside the decline in vaccine-targeted serotypes is referred to as serotype replacement (3, 4). A key question is how much each of the existing NVTs may be expected to expand in carriage prevalence as their competitors are removed by vaccination, and more generally, how the abundance of specific genetic lineages will change following vaccination.

Negative frequency-dependent selection (NFDS) on the accessory genome of *S. pneumoniae* is one evolutionary mechanism with some predictive power for post-vaccination strain dynamics in this species (5, 6). This mechanism posits that above a certain equilibrium frequency, the presence of a gene in a strain is deleterious, but below that frequency it is advantageous; therefore, the frequency of this gene tends to return to its equilibrium after vaccination perturbs the population. A model based on this concept allowed the dynamics of individual strains in the carriage population post-PCV7 to be modeled as a selective process in which NFDS changes strain frequencies to produce gene frequencies similar to those observed at the pre-vaccine equilibrium (6). The result was an ability to predict which strains would succeed, and which would not, following vaccination.

Pneumococcal serotypes have variable ability to cause IPD, which means that strains collected from population based IPD surveillance do not reflect how commonly different serotypes are carried. The pneumococci carried in the human nasopharynx are the populations that are transmitted from person to person and are therefore the bacterial populations upon which selection acts (7). Predictive or descriptive accounts of pneumococcal evolution therefore should be based on the population of colonizing pneumococci. However, in some circumstances much larger and more representative data sets are only available for pneumococci causing invasive disease; this is particularly true for the period post 2010 when the 13-valent pneumococcal conjugate vaccine (PCV13) was introduced in the United States.

The purpose of this study is therefore twofold. We first describe a simple weighting system that adjusts a sample of invasive disease isolates to better represent the carriage population, showing that this weighting system improves the fit of previously studied NFDS models. Then we use weighted invasive disease data to test whether such models can effectively predict the proportional distribution of pneumococcal colonizing clones in the post-PCV13 epoch.

## Results

### Weight values

An inverse measure of invasiveness as a weight was created to convert invasive disease abundance of isolates carrying each serotype to an estimate of the frequency of such isolates in carriage. From the pre-vaccine epoch, there were 6,976 specimens in the IPD data from 1998– 2000 and 384 isolates from 1998–2001 in a combined data set from the state of Massachusetts and from the Southwest United States. Using these data from the pre-vaccine epoch of 1998– 2000 in the United States, the weight for each serotype is the ratio of the proportion of the serotype in the carriage data to the proportion in the IPD data. The weight value for each serotype is displayed in Figure 1 (tabulated in Supplemental Data 1). The vaccine serotypes (VTs) in PCV7 and PCV13 typically had weight values lower than or close to 1, indicating that VTs were overrepresented in invasive disease compared to carriage. Many NVTs had weights above 1, indicating types that were more common in the carriage data. In the pre-vaccine epoch, 72% of the IPD population (31 serotypes including 8 VTs [4, 18C, 14, 6B, 9V, 7F, 1, 3] of which the last 3 were only in PCV13) had a weight <1, and 28% of the population (34 serotypes including 5 VTs [19F, 23F, 19A, 6A, 5] of which the last 3 were only in PCV13) had a weight >1.

**Figure 1.**
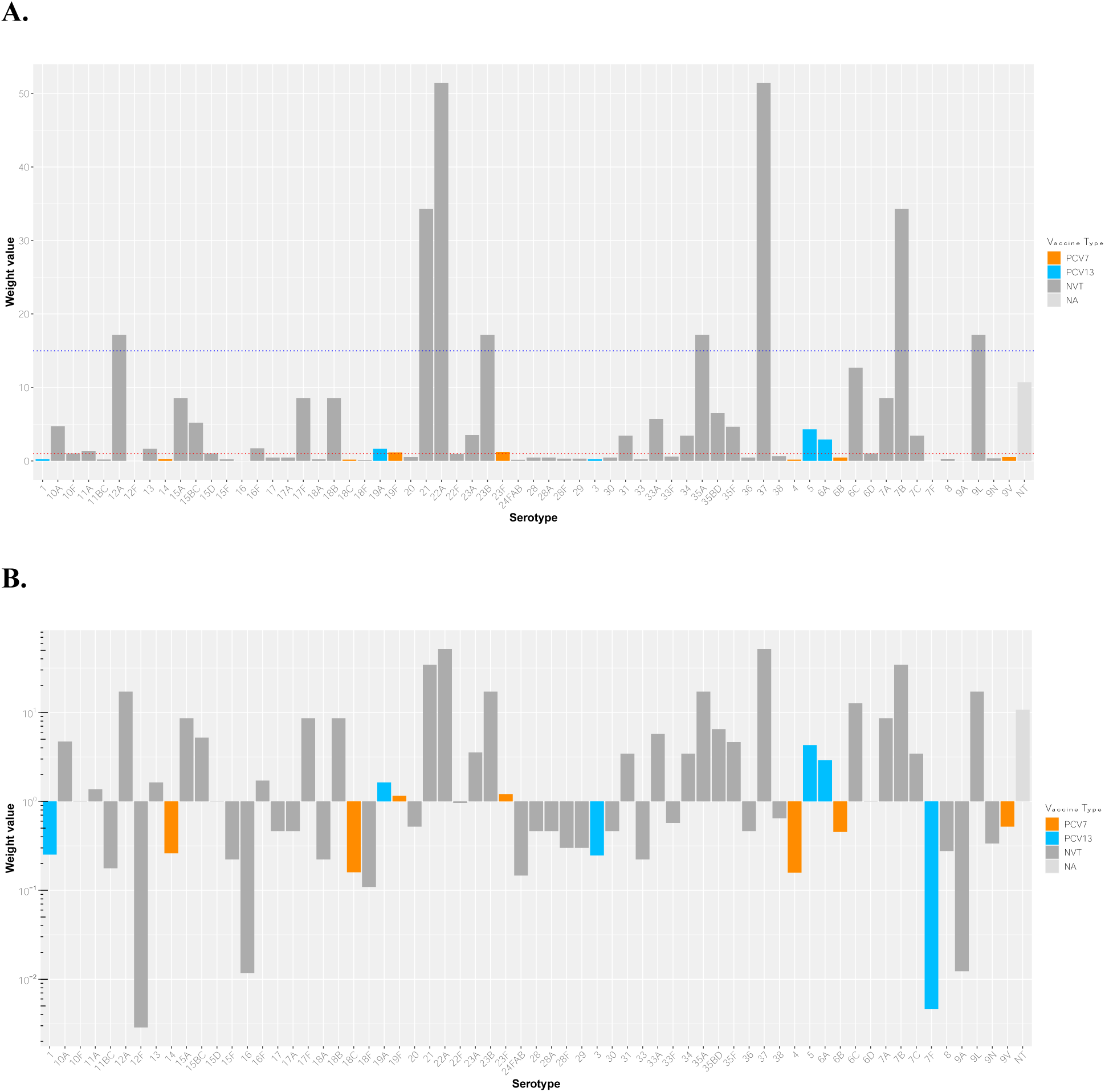
Weight value for each serotype. The weight was developed from the serotype distribution in the pre-vaccine epoch in the US (CDC ABCs IPD data from 1998–2000 and carriage data collected in Massachusetts and Southwest US from 1998–2001). There are 65 different serotypes, including the not-assigned (NA) group. **A. Linear scale.** The orange bars represent PCV7-covered serotypes, and the blue bars represent the additional six serotypes in PCV13. The red dotted line indicates the weight value is equal to 1, and the blue dotted line indicates the weight value is equal to 15. **B. Log scale**. These vaccine-covered serotypes generally had weight values less than or close to 1. Many non-vaccine types (NVTs) had weight values greater than 1. PCV7, 7-valent pneumococcal conjugate vaccine; PCV13, 13-valent pneumococcal conjugate vaccine; NVT, non-vaccine types; NA, unassigned serotype group.

Some serotypes had extreme weight values (≥15), but these made up a very small proportion of the IPD data (approximately 0.01%). There were 4 very small weight values (Figure 1B, weight < 0.01), including 12F, 16, 7F, and 9A, among which 7F is one of the PCV13 vaccine serotypes. These serotypes were only present in the IPD dataset; to get a hypothetical weight, we added a very small count for these serotypes in carriage data assuming that they were present at a negligible or non-detectable level. These 4 serotypes accounted for 6.4% of IPD isolates.

### Accessory gene frequency correlations before and after the application of weights

We assessed the effects of the weights applied to the IPD data by comparing the correlations in accessory gene frequency (i) between the carriage population and the invasive population and (ii) between the different time periods in invasive data, with and without weights. If weighting improved the representativeness of the IPD data for the carriage population, we would expect that these correlations would improve with the application of the weights. Indeed, the correlations of accessory gene frequencies between the carriage data and the IPD data in the linear scale were improved or in no case reduced for pre-vaccine (before weights: 0.92 vs. after weights: 0.94), post-PCV7 (0.91 vs. 0.93), and post-PCV13 (0.95 vs. 0.95) (Figure 2). The corresponding residual sum of squares was also reduced after applying the weights. Similar findings were obtained using logit-transformed frequencies where the weights improved or in no case reduced the correlation between the carriage data and the IPD data for pre-vaccine (before weight: 0.90 vs. after weight: 0.91), post-PCV7 (0.88 vs. 0.88), and post-PCV13 (0.89 vs. 0.91) (Supplemental Figure 1). The improved correlations suggest that the weights adjusted the invasive population to better reflect the carriage population.

**Figure 2.**
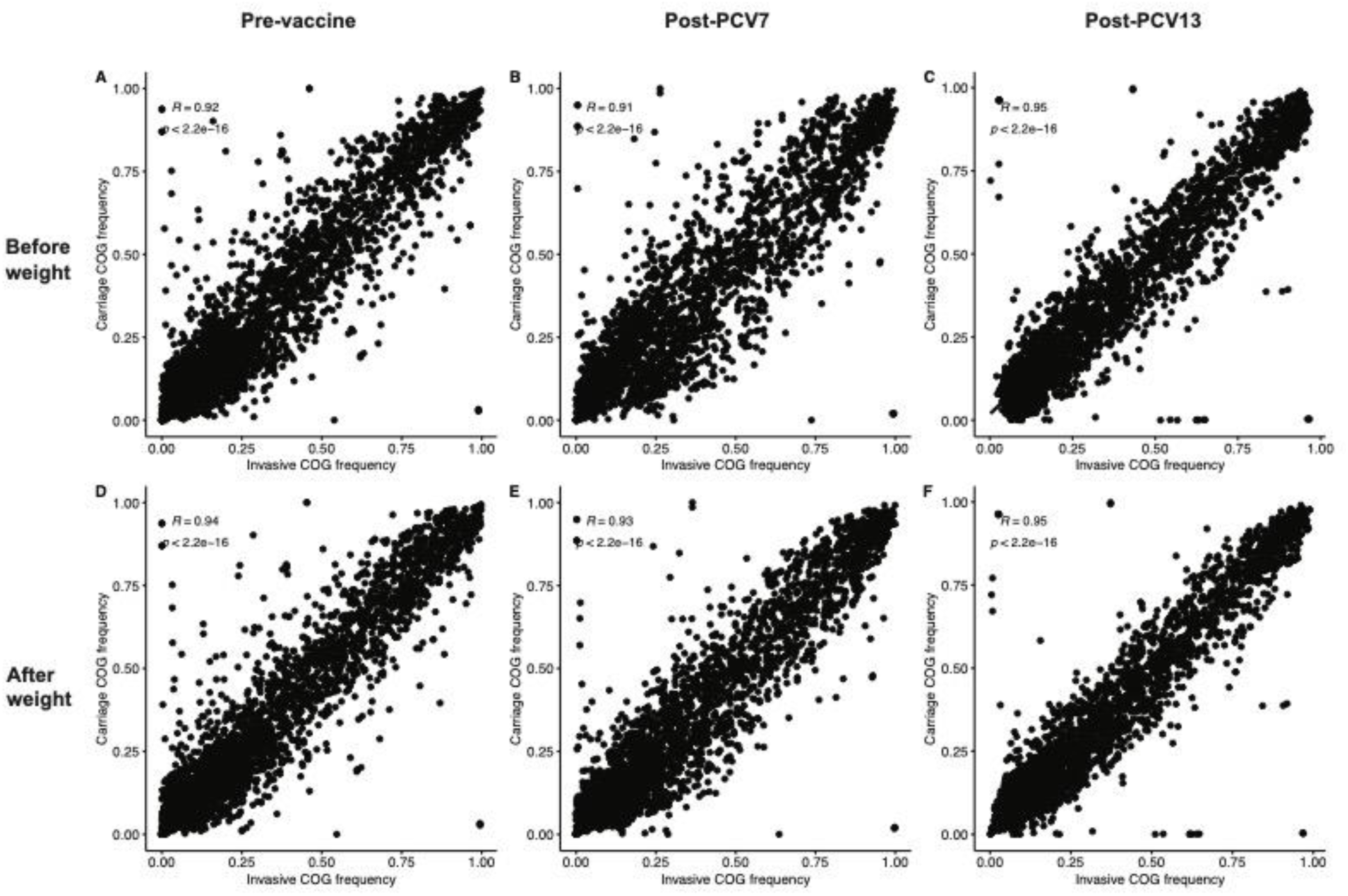
Correlations of accessory gene frequencies between the carriage data and the IPD data before and after weights. The upper panels A, B, and C represent the correlations between the carriage data and the IPD data before the application of weights for pre-vaccine (A), post-PCV7 (B), and post-PCV13 (C). The lower panels D, E, and F show the correlations for each epoch after the application of weights. We observed both visually tighter scatterplots and improved correlations after weights. COG, accessory clusters of orthologous genes; R, correlation coefficient; PCV7, 7-valent pneumococcal conjugate vaccine; PCV13, 13-valent pneumococcal conjugate vaccine.

Given the previous observations (8) in carriage data that accessory gene frequencies tended to return to the pre-vaccine equilibrium frequencies, we compared the correlations between populations before and after vaccination —pre-vaccine vs. post-PCV7, pre-vaccine vs. post-PCV13, and post-PCV7 vs. post-PCV13— in the IPD before (Figure 3A-C) and after applying the weights (Figure 3D-F). We observed that applying the weights to the IPD data improved the correlation of accessory gene frequencies between different epochs in the invasive data; similar information was revealed in the logit-transformed frequencies (Supplemental Figure 2).

**Figure 3.**
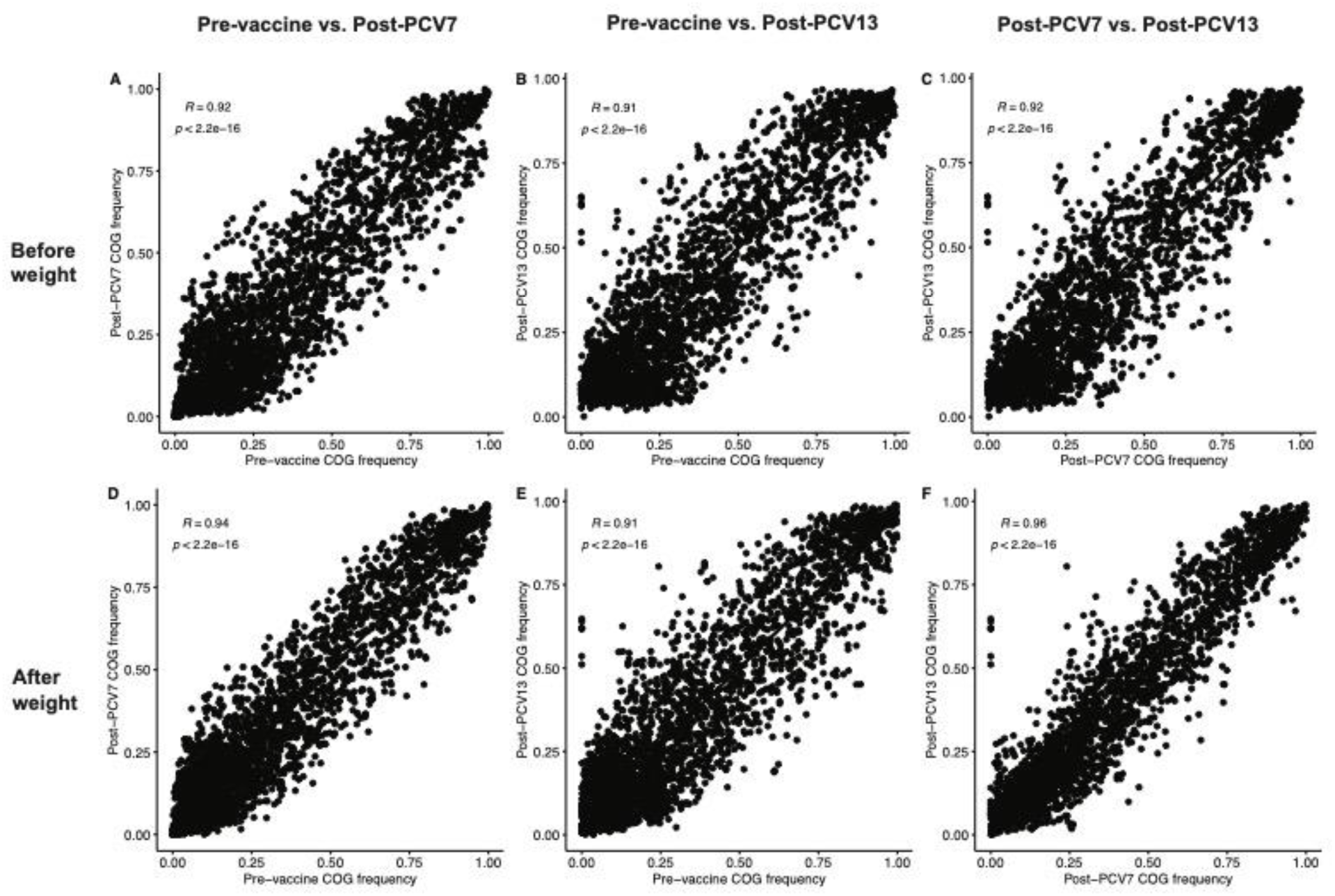
Correlations of accessory gene frequencies between different epochs in the IPD data before and after application of weights. A, B, C show the correlations before weights in the IPD data for pre-vaccine vs. post-PCV7 (A), pre-vaccine vs. post-PCV13 (B), and post-PCV7 vs. post-PCV13 (C). D, E, F show the correlations after weights in the IPD data for different epochs. We observed that the weights improved the correlations in the IPD data with both visually tighter scatterplots and improved correlation coefficients. COG, accessory clusters of orthologous genes; R, correlation coefficient; PCV7, 7-valent pneumococcal conjugate vaccine; PCV13, 13-valent pneumococcal conjugate vaccine.

### Post-PCV13 strain proportion prediction

To note, the term “strain” refers to the genetic cluster of closely related sequences identified by the population partitioning using nucleotide k-mers (PopPUNK) (45) based on the whole-genome assembly. Given the encouraging results obtained with improved accessory gene frequency correlations after the application of weights the NFDS model (6), was tested for predicting the post-PCV13 carriage population using IPD data. Because the collection contained many small sequence clusters as defined by the Global Pneumococcal Sequence Cluster (GPSC) (9) via PopPUNK, we combined the small GPSCs < 10 sequences by a recursive algorithm that combined any sequence cluster (SC) with <10 isolates with the most similar SC defined by the sum of absolute difference of each accessory clusters of orthologous gene (COG) frequency within the SC. Initially, 159 GPSCs were classified by PopPUNK, among which 109 GPSCs (n=212 isolates) contained ≤10 sequences. The combining process was repeated until all SC reached at least 10 sequences. We had four rounds of the combining process, and the number of GPSCs was reduced to 64. The sequence count in each strain after combined is shown in Supplemental Figure 4.

We further compared the observed strain proportions in the IPD data at E1 and E3 epochs, before (Figure 4A) and after (Figure 4B) weights applied, after combining the small GPSCs. The weights generally reduced the VTs in the strain and increased NVTs. The mixed strains could be increased or decreased depending on the proportion of vaccine/non-vaccine serotypes in a mixed strain.

**Figure 4.**
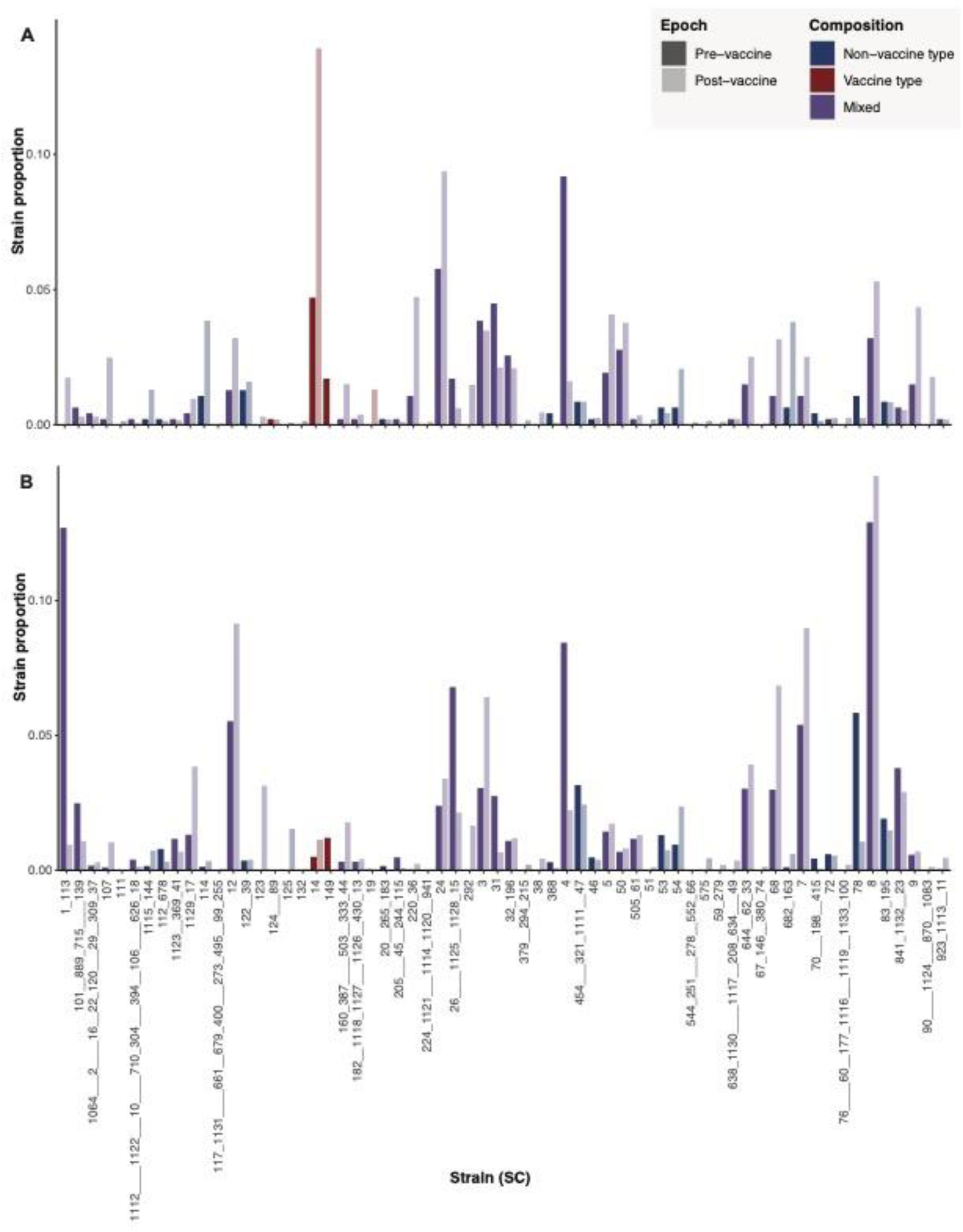
Strain proportion after combining the small GPSCs before and after application of weights. A is the strain proportion before the application of weights, and B is after. The name of the strain indicated the original GPSC called by PopPUNK. The underscore connection indicated the combining process by which GPSCs were combined together with minimal accessory frequency distance. The number of underscores indicated at which round out of the 4 rounds the two GPSCs were combined, for example, the connection by one underscore sign indicated that the two GPSCs were combined at the first round. The weights generally reduced the vaccine types in the strain and increased non-vaccine types. The mixed strains could be increased or decreased depending on the proportion of vaccine/non-vaccine serotypes in a mixed strain.

The NFDS model after weighting predicted the strain proportions in the projected carriage population better as compared with before weighting (Figure 5). The regression between the observed and predicted strain proportions was improved, with the adjusted R-squared increasing from 0.176 before weights to 0.544 after weights, indicating the impact of the bias introduced by the IPD sample on the power of the NFDS model; and the inverse-invasiveness weighting substantially improved the predictive power of the NFDS model. The regression coefficient between predicted and observed SC frequencies was 0.82 (95%CI: 0.62–1.01) after the application of weights. In addition, the sum of the squared prediction errors (SSE) was reduced after weighting. In the prediction model, outliers were defined as points for which the difference between the predicted and observed proportions was >1.5 times the interquartile range of the distribution of predicted and observed proportion differences. We found some outliers with a higher observed proportion than the predicted value, including SC-3, SC-7, and SC-12. SC-3 contained very diverse serotypes, mainly 9V (6.5%), 15A (25.7%), 15BC (15.3%), and 35BD (23.7%); SC-7 contained 38.8% of 23A and 60.1% of 23B; and the SC-12 contained 93.9% of serotype 15A. The outliers with higher predicted proportion than the observed included SC-4 (60.2% of 15BC and 37.7% of 19A), SC-26_1125_1128_15 (78.3% of 6C), and SC-78 (94.8% of 6C). These outliers may be related to the potential cross-reactivity of their serotypes with VTs or an expanded ecological niche after the vaccine types were removed.

**Figure 5.**
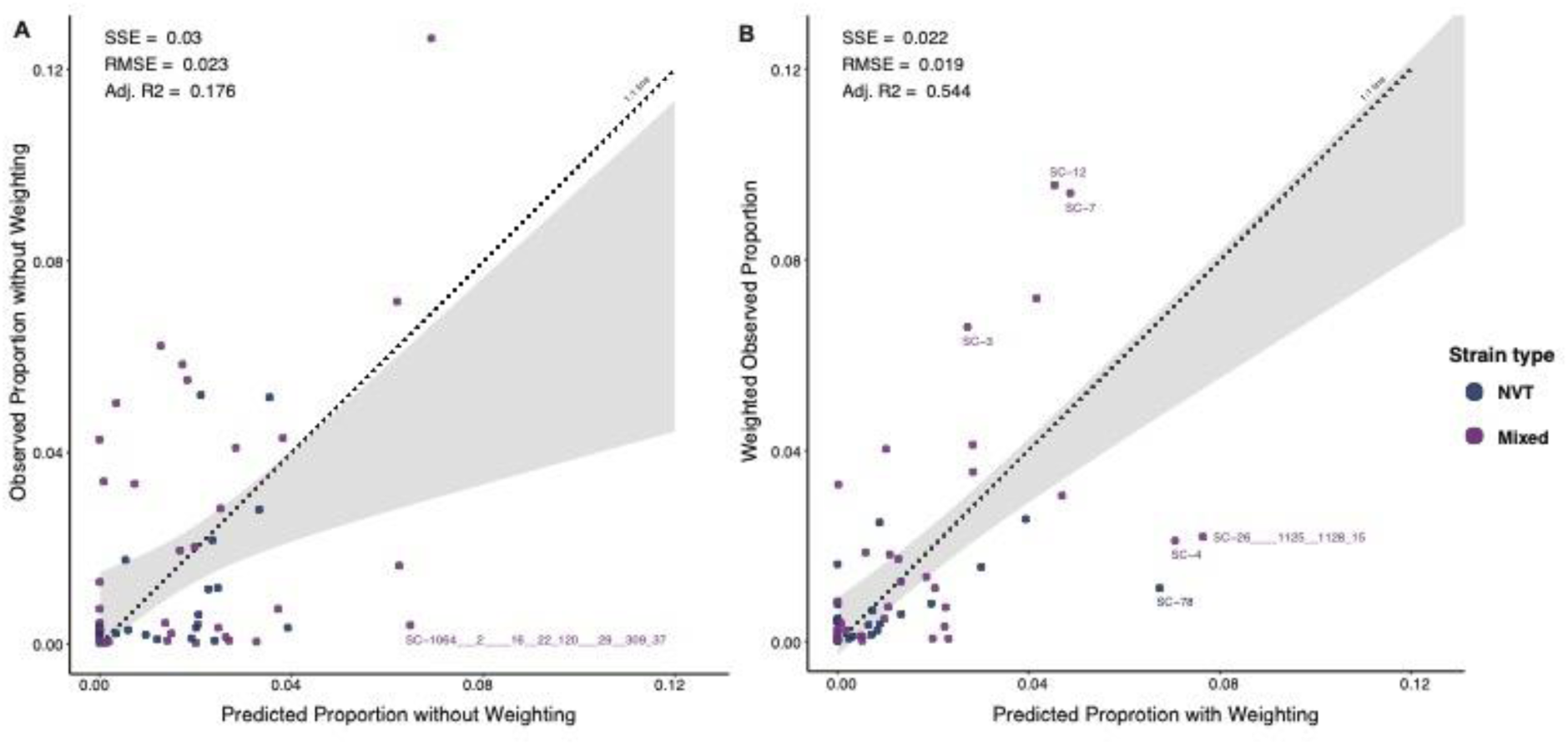
The NFDS model predicted strain proportions before and after the application of weights. Scatterplot of observed versus predicted proportions of 58 strains at post-PCV13 equilibrium is based on quadratic programming. These 58 strains contained at least 1 NVT isolate pre-vaccine or were imputed with one isolate pre-vaccine. Perfect predictions fall on the dotted line of equality (1:1 line). The shaded gray region shows the confidence interval from the linear regression model used to test for deviation of the observed versus predicted values compared with the 1:1 line. A is the prediction based on the IPD data before the application of weights, and B is after weights. Outliers are defined as the difference between their predicted and observed proportion that is >1.5 times the interquartile range of the distribution of predicted and observed proportion differences. The outliers with higher observed proportion than the predicted are SC-3, SC-12 and SC-7. The SC-3 contained very diverse serotypes, mainly 9V (6.5%), 15A (25.7%), 15BC (15.3%), and 35BD (23.7%); the SC-7 contained 38.8% of 23A and 60.1% of 23B; and the SC-12 contained 93.9% of serotype 15A. The outliers with higher predicted proportion than the observed included SC-4 (60.2% of 15BC and 37.7% of 19A), SC-26___1125__1128_15 (78.4% of 6C), and SC-78 (94.8% of 6C). NVT, non-vaccine type; Mixed, strain containing both vaccine type and non-vaccine type; SC, sequence cluster or strain; SSE, sum of squares due to error; RMSE, root mean square error; Adj. R2, adjusted R-squared; NFDS, negative frequency-dependent selection; PCV13, 13-valent pneumococcal conjugate vaccine.

### Sensitivity analysis

Epidemiological surveillance showed that PCV13 did not decrease the carriage prevalence of serotype 3 after vaccine introduction even though serotype 3 was a component of the vaccine; therefore, in a separate sensitivity analysis, we repeated the above but treated serotype 3 as an NVT. Serotype 3 had 1,453 sequences (12.3% before weights) in the IPD data. The NFDS model substantially improved the prediction accuracy after weights (adjusted R-squared increased from 0.23 before the application of weights to 0.55 afterward). The adjusted R-squared was generally improved when compared with treating serotype 3 as VT. The regression coefficient between predicted and observed strain proportions was very similar to treating serotype 3 as a VT, that is, 0.81 (95%CI: 0.62–1.00) after the application of weights. The outliers after weighting when treating serotype 3 as NVT were the same as treating serotype 3 as VT. However, two outliers were reported before weighting when treating serotype 3 as an NVT: SC-14 (99.9% of serotype 3) and SC-1_113 (92.8% of serotype 19A and 5% of 19F).

## Discussion

To predict the population dynamics of *S. pneumoniae* after PCV13 introduction and work around the limited availability of carriage data with samples collected over the required period, we developed a weighting system based on the distribution of serotypes in the carriage data and invasive disease data in the United States during the pre-vaccine epoch (i.e., carrier-to-case ratio for each serotype). The weighting adjusted the CDC ABCs invasive disease dataset to approximate the strain composition of the carriage population, resulting in improved correlations of accessory gene frequencies between the carriage data and invasive data after weights. With the weights applied, the NFDS model acquired predictive power for strain proportions in the carriage population in the post-PCV13 epoch. After weighting, the NFDS model consistently demonstrated improved performance, which confirmed the value of the weight system. This was further corroborated by the sensitivity analyses treating serotype 3 as a NVT. Therefore, our weight system shows how genomic sequences from invasive disease surveillance can be used to study evolutionary mechanisms and predict pneumococcal population dynamics in carriage, even where invasive disease reflects a biased sample of the carriage population.

The weight value estimates the inverse of the invasiveness of each serotype based on the *S. pneumoniae* population in the United States. Previous studies have quantified the tendency of different serotypes to cause invasive disease by the case-to-carrier ratio or invasiveness index (10, 11). The major determinant of invasiveness is the capsular serotype, and most serotypes have known invasiveness estimates from epidemiological or experimental data (10, 12–14). Therefore, invasive disease datasets can reflect the carriage population, but with the more invasive serotypes overrepresented. We used a composite data set of a carriage study primarily in children <5 and one in children <7 years old, and an all-ages invasive disease data set to arrive at our weights. This represents a working hypothesis that carriage in young children is the primary population for pneumococcal transmission and evolution, consistent with evidence that young children are key drivers of pneumococcal population dynamics. Several lines of evidence suggest this role, most notably the substantial drop in IPD in all ages following infant and toddler vaccination with PCV7 (15, 16). The use of an all-ages invasive disease data set, ABCs, to develop the weights was appropriate because it was this data set which was used for NFDS model predictions, and the purpose of the weights is to transform a given invasive disease data set to frequencies more representative of carriage in the population that is undergoing evolutionary pressure. In addition, the weight was calculated based on the serotype distribution during the pre-vaccine epoch during 1998–2001. This is a deliberate methodological choice because the pre-vaccine serotype-specific weight represents when the population was under equilibrium and not perturbed by the PCVs, serving the purpose of performing the NFDS prediction model. But it can be one of the reasons reducing the model’s fit to the data because in several instances the clonal compositions of serotypes changed between pre-PCV7 and subsequent epochs. This imposes a limitation of serotype-specific weighting. A potentially more sophisticated approach is to compute a strain-specific weight considering the serotype component change before and after vaccination.

PCVs have been effective at reducing pneumococcal disease worldwide. However, replacement with non-PCV serotypes remains a concern, such as the increase of serotype 19A after PCV7 introduction in the United States in 2000 (17, 18). After PCV13 introduction, replacement was also observed in the United Kingdom (with the important proviso that serotype 3 remained a consistent cause of IPD) (19–21). In the prediction model our sensitivity analysis, treating serotype 3 as NVT, generated a more accurate prediction of the strain proportion post-PCV13. We also observed some outliers, SCs which made up a proportion of the equilibrium population that was either substantially more or less than predicted. When examining the components of serotypes in these strains, we found the strains (SC-4, 15, and 78) that were overestimated by the NFDS model were associated with serotypes 15BC and 6C; in the case of 6C this could represent partial cross protection against 6C from the 6A and 6B components of PCV13 (22). We also found that serotypes 23A, 23B, and 15A were common among those the model underestimated (SC-7 and 12), suggesting that these serotypes had additional advantages compared with other NVTs. One explanation may be that the proportion of penicillin-nonsusceptible clones in 15A, 23A, and 23B have dramatically increased post-PCV7, and this has continued into post-PCV13 epoch (23). Although not observed in the United States, an increase in serotype 24F caused a sizeable increase in pneumococcal meningitis cases in many countries after 5 years of the PCV13 rollout (24). The advantage of serotype 24F, belonging to GPSC10, was potentially driven by multidrug resistance and lack of immunity in the host population (24). Hence, to fully explain the response of the pneumococcal population after introduction of successive PCVs in the United States, other mechanisms should be investigated with continued pneumococcal invasive disease or carriage population surveillance.

There are some limitations in this study. First, sampling biases in host age and/or geographic regions existed in the carriage data and the invasive disease data, resulting in inaccuracy in the weight system. Although the CDC ABCs invasive disease data collected from ten states/regions across the United States were reasonably representative of the population diversity, the carriage data were collected from Massachusetts and the Southwest United States and may not have fully represented the serotype and strain diversity across the entire United States. Additional noise may have arisen from errors in serotype identification. Although the whole-genome approach to identifying the sample serotypes was highly consistent with laboratory identification, some errors or ambiguous assignments occurred (25, 26). Second, weights developed from the U.S. data may not work for *S. pneumoniae* populations in other locations because of spatial heterogeneity in the bacterial population. For example, in the carriage population from Maela, Thailand during 2001–2007 in the absence of vaccination, the serotype distribution and strain diversity was very different from the pre-vaccine population from other locations (5, 27). However, our weight values were highly correlated with the weights developed using epidemiological data from different global regions, suggesting these values could be used for other locations, but interpretation should take the heterogeneity into account. Third, in the NFDS model, we assumed that after 7 years of the PCV13 rollout, the pneumococcal carriage population had reached post-PCV13 equilibrium. However, previous evidence has shown that the population took approximately 10 years after PCV7 to reach equilibrium (8). As the CDC ABCs is ongoing, more data will become available for further testing the model.

In conclusion, the weight system effectively adjusted invasive disease surveillance data to better represent the genomic components of the carriage population of *S. pneumoniae*. Weighting allowed us to predict the carriage strain proportion in the population in the post-PCV13 epoch, although other mechanisms should be further investigated to explain certain emerging strains after the rollout of PCVs. Our methods have enriched the value of genomic sequences from invasive disease surveillance that is readily available, easy to collect, and of direct interest to public health.

## Methods

### Study population

We used three sources of data: i) carriage data from children aged <7 years in Massachusetts, US (hereafter referred to as MA data) (28, 29), ii) carriage data from Native American communities in the Southwest US (hereafter referred to as Southwest US data) (30–32), and iii) invasive pneumococcal disease data from CDC ABCs (hereafter referred to as IPD data) (33–36).

In the MA data, pneumococcal isolates were collected from nasopharyngeal swabs of children under the age of 7 years at a participating primary care provider in communities throughout Massachusetts (28, 29). Samples were collected between October and April of 2000–2001, 2003–2004, 2006–2007, 2008–2009, 2010–2011, and 2013–2014. This final dataset contained 1,314 genomic sequences and associated metadata.

The Southwest US data included pneumococci isolated from a subset of participants in three prospective, observational cohort studies of pneumococcal carriage among southwest Native American individuals from 1998 through 2012 described elsewhere (6, 30–32). Briefly, participants living on Tribal lands in the Southwest US were enrolled during three periods: 1998–2001, 2006–2008, and 2010–2012. Nasopharyngeal swab specimens were obtained during visits to healthcare facilities or the participant homes to determine pneumococcal carriage status. Except for a subset of isolates collected from 2006–2008, all isolates were obtained from children <5 years. The Southwest US data included 937 genomic sequences and associated metadata that were previously published using a random selection of isolates from each time period, with an oversampling of isolates post-PCV7.

All the IPD data from 1998 through 2018 were identified through CDC ABCs, an active population- and laboratory-based system that covers a population of approximately 34.5 million individuals in ten geographic locations in the US, including California, Colorado, Connecticut, Georgia, Maryland, Minnesota, New Mexico, New York State, Oregon, and Tennessee. ABCs areas, methods, and key surveillance data through 2018 are described elsewhere (34–36). Briefly, ABCs personnel routinely contacted all microbiology laboratories serving acute care hospitals in their area to identify cases. Standardized case report forms that included information on demographic characteristics, clinical syndrome, and outcome of illness were completed for each identified case. Whole genome sequencing-based characterization was conducted on all pneumococcal isolates, which included identifying capsular serotype and predicting minimum inhibitory concentration (MIC). A case of IPD was defined as *S. pneumoniae* detection from a normally sterile site in a surveillance area resident; common IPD syndromes included bacteremia with pneumonia, bacteremia without focus, and meningitis. Isolates were assigned serotypes at CDC’s Streptococcus Laboratory or the Minnesota Department of Health using Quellung (1998–2015) and whole-genome sequencing (WGS; 2015–2018) (37). The final IPD dataset had 11,795 sequences and associated metadata.

### Epoch definition

Epochs were defined to determined which time periods were used to generate weights and which were used to model different periods of vaccination. To generate weights, the MA data from 2000–2001 were defined as pre-vaccine epoch (E1) because PCV7 started roll-out for children in 2000, and we considered that this vaccine had not yet perturbed the bacterial population; 2003– 2009 as post-PCV7 epoch (E2); and 2010–2014 as post-PCV13 epoch (E3). With the Southwest US data, we defined 1998–2001, 2006–2008, and 2010–2012 as E1, E2, and E3, respectively. With the IPD data, for the weight development, we defined 1998–2000 as E1, 2009 as E2, and 2013–2018 as E3. For predicting post-PCV13 population dynamics, we defined 2009 as the pre-vaccine period for PCV13 (Pre-PCV13 E1), assuming that the population had reached a new equilibrium post-PCV7; 2013–2016 as Post-PCV13 E2, the removal stage of vaccine types in the population; and 2017–2018 as Post-PCV13 E3, assuming that the new equilibrium post-PCV13 had been reached. In this study, the NFDS model attempted to predict the strain proportions in 2017–2018, as this was the longest time period after the introduction of PCV13 for which data were available.

### Serotype-specific Weights

Invasiveness measures for pneumococcal serotypes have been defined in various ways, each monotonically associated with the ratio of invasive disease incidence to carriage prevalence for that serotype in a particular population (10, 38). While other determinants may play a role in invasiveness, serotype accounts for much of the variation in this property (10–12, 14). We therefore chose to use an inverse measure of invasiveness as a weight to convert invasive disease abundance of isolates carrying each serotype to an estimate of the frequency of such isolates in carriage. Specifically, using data from the pre-vaccine epoch of 1998–2000 in the US, the weight for each serotype is the ratio of the proportion of the serotype in the carriage data to the proportion in the IPD data. We used all cases in the ABCs data from the pre-vaccine period, whether the isolate from the case was sequenced or not. For serotypes that were only present in the IPD data, we added a small count (n=0.05) to each of these serotypes in the carriage data to generate an approximate weight value. For those serotypes that were only present in the carriage data, we did not calculate a weight. For the serotypes that were neither in the IPD pre-vaccine epoch nor in the carriage data but emerged after the vaccine was introduced, we applied a very small count (0.05 for each of the serotypes in the carriage data and proportionally 0.85 for the IPD data) to generate a hypothetical weight that was close to 1. Some ambiguous serotypes, for which inference from whole-genome sequence was not possible, were assigned to a most likely serotype as described in Supplemental Methods 1.

### Bioinformatics pipeline

All genomic and associated epidemiological data are publicly available (8, 28, 29, 35, 36) (National Center for Biotechnology Information NCBI accession numbers provided in Supplemental Data 2). Raw sequence reads were downloaded from the Sequence Read Archive (SRA) in the NCBI database. Genomic assembly from raw reads was constructed using SPAdes v3.10 (39) and annotated using Prokka v1.11 (40) embedded in Unicycler (41). Quast v4.4 (42, 43) was used to assess the quality of each genomic assembly. A sequence assembly was excluded when it had i) an N50 less than 15 kb; or ii) ≥500 contigs, indicating the genome was too segmented; or iii) a genome length <1.9 Mb or > 2.4 Mb; or iv) a Global Pneumococcal Sequence Cluster (GPSC) (9) not assigned due to “genomes with outlying lengths detected.” With the reference genome ATCC 700669 (5), Roary v3.10.0 (44) was then used to identify core and accessory genes. The core genes were defined as being present in >99% of the isolates to generate a core gene alignment, whereas accessory genes were present in 5% to 95% of the isolates and formed into a matrix of absence (value of 0) and presence (value of 1) for each sample. There were 3,166 accessory genes identified in the IPD population.

### Strain definition

The genetically similar sequence clusters (SCs)—strains—were defined by the population partitioning using nucleotide k-mers (PopPUNK) (45) based on the whole-genome assembly. The PopPUNK approach followed instructions from the Global Pneumococcal Sequencing project (https://www.pneumogen.net/gps/assigningGPSCs.html). The advantages of SCs defined by PopPUNK are that they have a global definition and can be compared across different populations. However, the major challenge posed by the approach is that many SCs contain one sequence or just a few sequences which lost the power of prediction due to limited information in the SC, resulting in the removal of these SCs to conduct the prediction for post-PCV13 epoch. To avoid losing data, we developed an accessory gene-based approach to combine the small SCs.

Specifically, SCs with ≤10 sequences were the targets to be combined. The approach accounted for accessory gene presence/absence distance between any of two SCs. The goal was to combine two SCs with the closest accessory gene presence/absence similarity. Under each SC, each accessory gene was calculated with proportion based on the presence/absence matrix. Then the sum of absolute difference of each accessory gene proportion was calculated. The SCs with ≤ 10 sequences were combined with their most similar SCs based on the accessory gene presence/absence distance. We repeated the process until all SCs contained ≥ 10 sequences.

### Weights

To adjust the accessory gene component of the invasive disease population to represent carriage, we applied the weights to the invasive accessory gene matrix of CDC ABCs that were collected and sequenced from 1998–2018. This was done by directly multiplying these weights by the accessory gene matrix of the absence (value of 0) and presence (value of 1). Calculation of accessory gene frequency before and after weights was demonstrated in Supplemental Methods 2. We assessed the effect of the weights cross-sectionally with the correlation of accessory gene frequencies between the carriage data and the invasive data before and after weights, and longitudinally with the correlation between pre- and post-vaccine in the invasive data before and after weights. The correlation coefficient and the residual sum of squares (RSS) were used to measure whether the correlations of accessory gene frequencies were improved before and after weights. To reduce heteroskedasticity, we repeated these analyses with logit-transformed frequencies. The analysis was conducted in RStudio v1.0.143 with R v4.1.2.

To adjust strain distribution in the invasive disease data to represent carriage, we calculated strain proportions based on serotype-specific weights, that is, the weights were applied to adjust the serotype components in each strain. Calculations of strain proportion before and after weights were demonstrated in Supplemental Methods 2.

### NFDS prediction model

Using the 3,166 accessory genes present in 5%–95% of taxa in the IPD data, pre-vaccine accessory gene frequencies were calculated for each strain, focusing on NVT taxa only. We considered strains that i) had NVT taxa present pre-vaccine and ii) were not too small if assigned by PopPUNK with whole-genome assembly. The later was achieved by combining small SCs as described above under “strain definition”. We applied the weights to the invasive accessory gene matrix of CDC ABCs that were collected and sequenced from 1998–2018 by directly multiplying these weights by the accessory gene matrix of the absence (value of 0) and presence (value of 1) to adjust the COG frequency. In addition, to adjust the observed strain proportion, the weights were applied to adjust the serotype components in each strain (Supplemental Methods 2). We then computed the predicted proportion of each strain such that post-vaccine accessory gene frequencies approached pre-vaccine frequencies as closely as possible by using a quadratic programming approach constructed using the package quadprog v1.5–5 implemented in RStudio v1.0.143 with R v4.1.2 (6). Details of this implementation can be found in the R code provided (https://github.com/c2-d2/Predicting_Pneumo_postPCV13). Some SCs had no presence in the pre-vaccine (Pre-PSV13 E1) epoch but emerged in the post-vaccine (Post-PCV13 E2 or E3) epochs: if one SC emerged in Post-PCV13 E2 and E3, then COG frequencies from Post-PCV13 E2 were used as the pre-vaccine equilibrium frequency with the assumption that one isolate in the pre-vaccine had its strain frequency; and if one SC emerged only in Post-PCV13 E3, then COG frequencies from Post-PCV13 E3 were used as the pre-vaccine equilibrium frequency with assuming one isolate in the pre-vaccine to impute its strain frequency. In our dataset, the majority of imputed SCs used COG frequencies from Post-PCV13 E2 as the proxy.

We then conducted the NFDS prediction models before and after weighting. For each model, accuracy would be reflected by a slope close to one and an intercept close to zero in the regression between the predicted and observed strain proportions. Goodness-of-fit statistics including sum of squares due to error (SSE), root mean squared error (RMSE), and degrees of freedom adjusted R-squared (Adj. R2) were used to evaluate each model.

### Sensitivity analysis

Epidemiological surveillance showed that the serotype 3 in PCV13 did not decrease the prevalence of this serotype over the years after the vaccine was introduced (19–21). Thus, in a separate sensitivity analysis, serotype 3 was treated as an NVT to recalculate the strains that had NVT taxa present pre-vaccine and conducted the prediction model.

## Data availability

Whole-genome sequencing data are publicly available before the initiation of the study in NCBI under BioProject number PRJEB2632 (https://www.ncbi.nlm.nih.gov/bioproject/PRJEB2632), PRJEB8327(https://www.ncbi.nlm.nih.gov/bioproject/PRJEB8327) and PRJNA284954 (https://www.ncbi.nlm.nih.gov/bioproject/PRJNA284954). Accession numbers and accompanying metadata have previously been published. The list of NCBI accession numbers for used sequencing data and all R scripts to perform weighting and prediction can be found in GitHub Repository (https://github.com/c2-d2/Predicting_Pneumo_postPCV13).

## ACKNOWLEDGMENTS

XQ and ML thank the funding support from the Waking Up, Lantern Ventures, the Morris-Singer Fund, and Award Number U54GM088558 from the National Institute of General Medical Sciences. The content is solely the responsibility of the authors and does not necessarily represent the official views of the National Institute of General Medical Sciences or the National Institutes of Health.

## Supplemental materials

**Supplemental Methods 1. Developing serotype-specific weight for ambiguous serotypes**

Some of the genome-assigned capsular serotypes were ambiguous. We made decisions for each specific scenario. We combined some serotypes together to remove the impact of the ambiguous serotypes. For example, 15B, 15C, and 15B/C were combined as 15BC to have one weight calculated rather than a weight for each. For other ambiguous serotypes, it was unreasonable to combine. For example, three isolates marked as serotype 18B/18C were in the IPD data before 2000; 18C with a weight value of 0.15 was covered by PCV7 and 18B with a weight value of 8.57 was NVT; thus, serotypes 18B and 18C were not combined. Instead, given the very high probability of an invasive case being 18C, these three isolates of 18B/18C were reassigned to 18C. There was one isolate marked as 7A/7F in the IPD in 2009. Serotype 7F was not in the carriage data in the pre-vaccine epoch, but there were 2.6% (n=184) 7F and 0.03% (n=2) 7A in the IPD pre-vaccine epoch; and 7F was covered in the PCV13. Therefore, considering the high probability of an IPD case caused by 7F, the one 7A/7F isolate was reassigned as 7F.

**Supplemental Methods 2. Demonstration of how weights were applied**

The table below shows a hypothetical data frame. Taxa mean each sequenced specimen; strain is the PopPUNK-defined sequence cluster (SC); serotype is a serologically distinctive group; weight is developed based on the serotype-specific inverse-invasiveness; and accessory clusters of orthologous gene COG1 to COGi are the accessory genes presenting in 5%-95% of all the IPD isolates.

To adjust strain distribution in the invasive disease data to represent carriage, we calculated strain proportion based on serotype-specific weights, that is, the weights were applied to adjust the serotype components in each strain. For example, SC-1 proportion is 4/7 (4 isolates of SC-1 over the total population size 7) before weights applied and (2*w1 + 2*w2)/(2*w1 + 3*w2 + 2*w3) after weights, i.e., the sum of weights for SC-1 over the sum of weights in the overall population. Similarly, SC-2 proportion is 3/7 before weights applied and (w2 + 2*w3)/(2*w1 + 3*w2 + 2*w3) after weights.

To adjust the accessory gene component of the invasive disease population to represent carriage, we applied the weights to the invasive COG matrix by directly multiplying these weights by the accessory gene matrix of the absence (value of 0) and presence (value of 1). For example, COG1 frequency in the overall population is 4/7 before weights applied and (w1 + w2 + 2*w3)/(2*w1 + 3*w2 + 2*w3) after weights, that is, the sum of weights when COG1 presents over the sum of all weights in the population. When calculating strain-specific COG frequency for prediction, the table was broken down by strains. For example, SC-1 COG1 frequency is ¼ before weights and w1/(2*w1+2*w2) after weights.

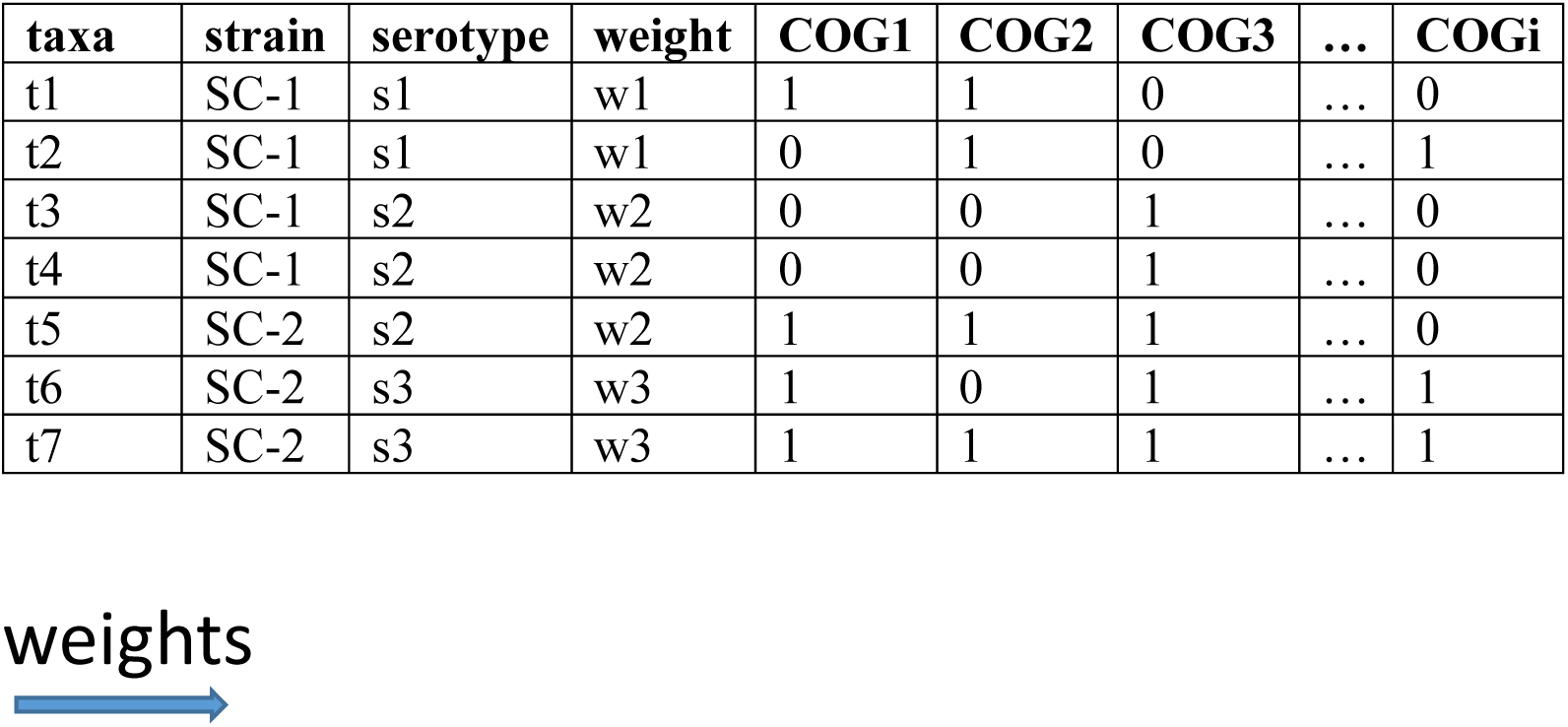
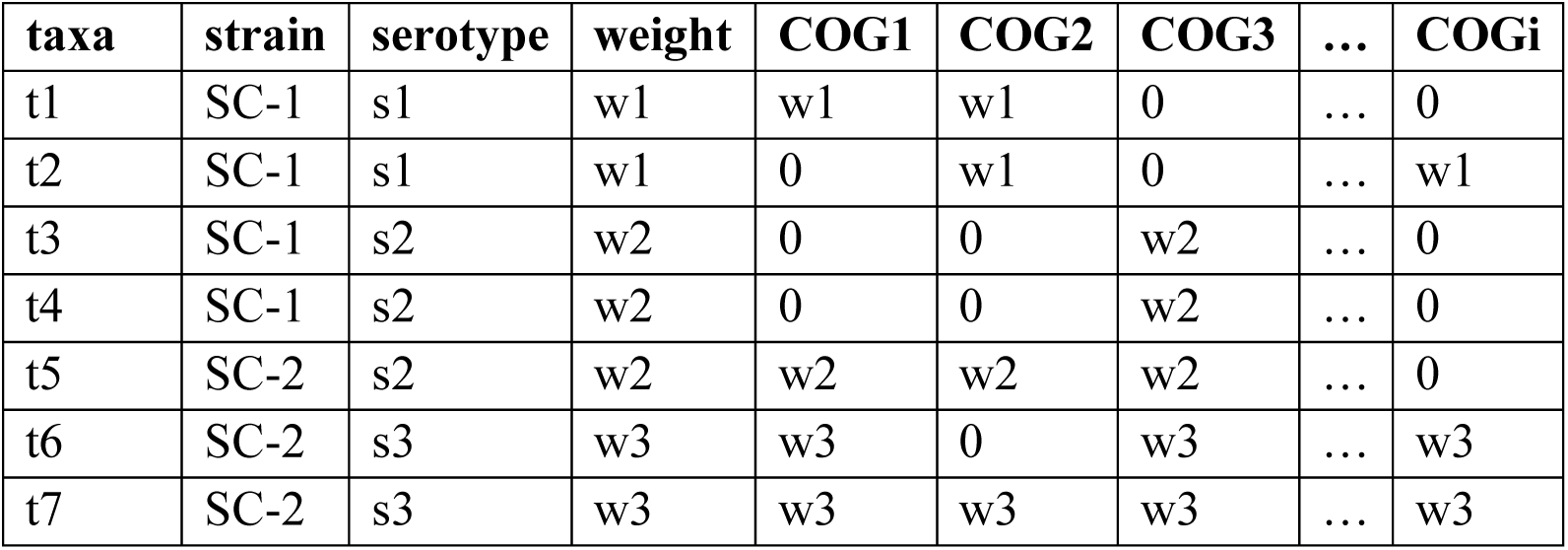

| taxa | strain | serotype | weight | COG1 | COG2 | COG3 | ... | COGi |
| --- | --- | --- | --- | --- | --- | --- | --- | --- |
| t1 | SC-1 | s1 | w1 | 1 | 1 | 0 | ... | 0 |
| t2 | SC-1 | s1 | w1 | 0 | 1 | 0 | ... | 1 |
| t3 | SC-1 | s2 | w2 | 0 | 0 | 1 | ... | 0 |
| t4 | SC-1 | s2 | w2 | 0 | 0 | 1 | ... | 0 |
| t5 | SC-2 | s2 | w2 | 1 | 1 | 1 | ... | 0 |
| t6 | SC-2 | s3 | w3 | 1 | 0 | 1 | ... | 1 |
| t7 | SC-2 | s3 | w3 | 1 | 1 | 1 | ... | 1 |
| t1 | SC-1 | s1 | w1 | w1 | w1 | 0 | ... | 0 |
| t2 | SC-1 | s1 | w1 | 0 | w1 | 0 | ... | w1 |
| t3 | SC-1 | s2 | w2 | 0 | 0 | w2 | ... | 0 |
| t4 | SC-1 | s2 | w2 | 0 | 0 | w2 | ... | 0 |
| t5 | SC-2 | s2 | w2 | w2 | w2 | w2 | ... | 0 |
| t6 | SC-2 | s3 | w3 | w3 | 0 | w3 | ... | w3 |
| t7 | SC-2 | s3 | w3 | w3 | w3 | w3 | ... | w3 |

**Supplemental Data 1. Weight values for each serotype** (https://github.com/c2-d2/Predicting_Pneumo_postPCV13)

**Supplemental Data 2. GenBank accession numbers for all genomic data used in this study (**https://github.com/c2-d2/Predicting_Pneumo_postPCV13)

---

**Supplemental Figure 1.**
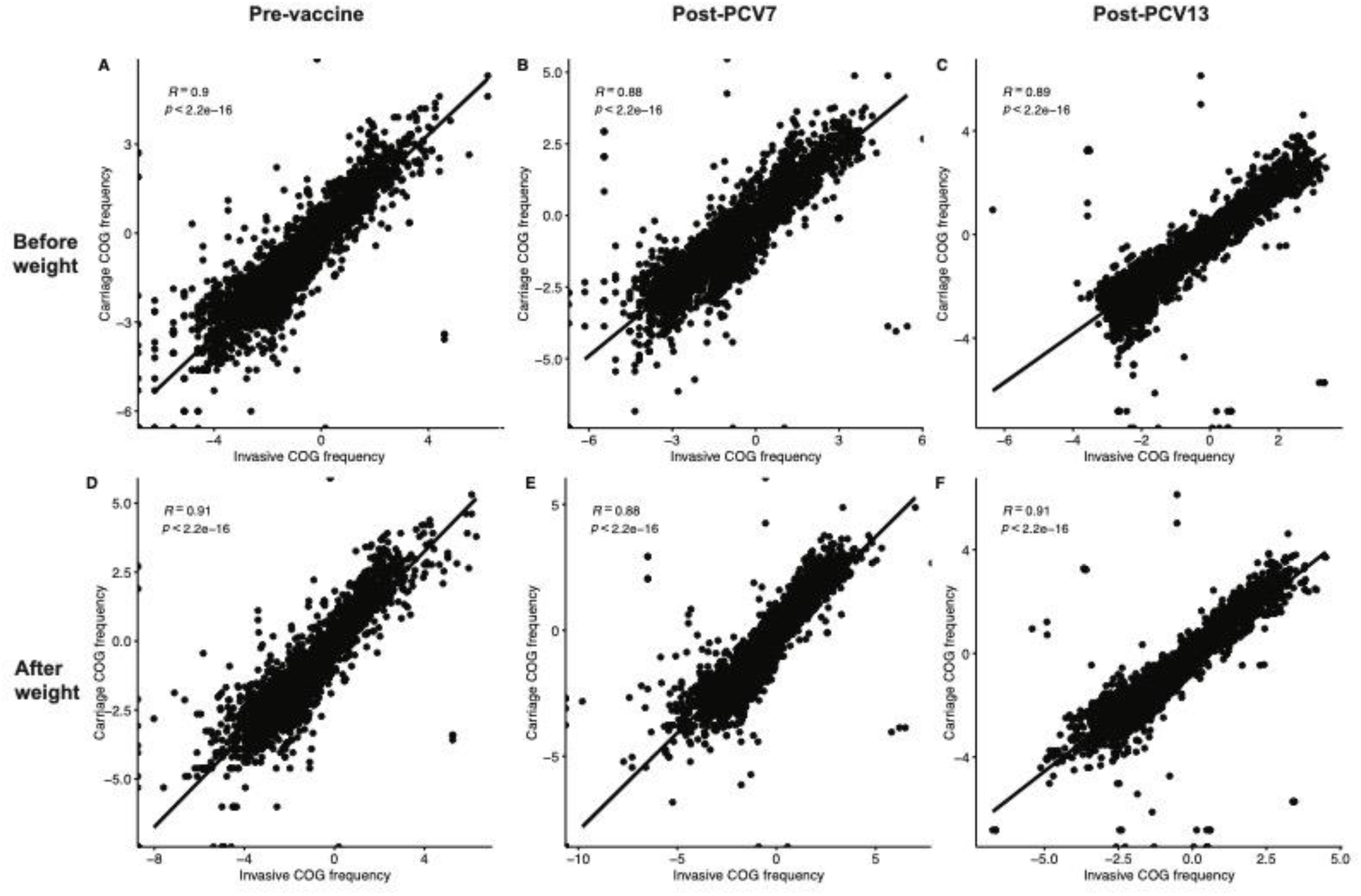
Correlation of logit-transformed accessory gene frequency between carriage and invasive data before and after application of weights. The upper panels A, B, and C show the correlations between the carriage data and the IPD data before the application of weights for pre-vaccine (A), post-PCV7 (B), and post-PCV13 (C). The lower panels D, E, and F show the correlations for each epoch after the application of weights. We observed both visually tighter scatterplots and improved correlations after weights. COG, accessory clusters of orthologous genes; R, correlation coefficient; PCV7, 7-valent pneumococcal conjugate vaccine; PCV13, 13-valent pneumococcal conjugate vaccine.

**Supplemental Figure 2.**
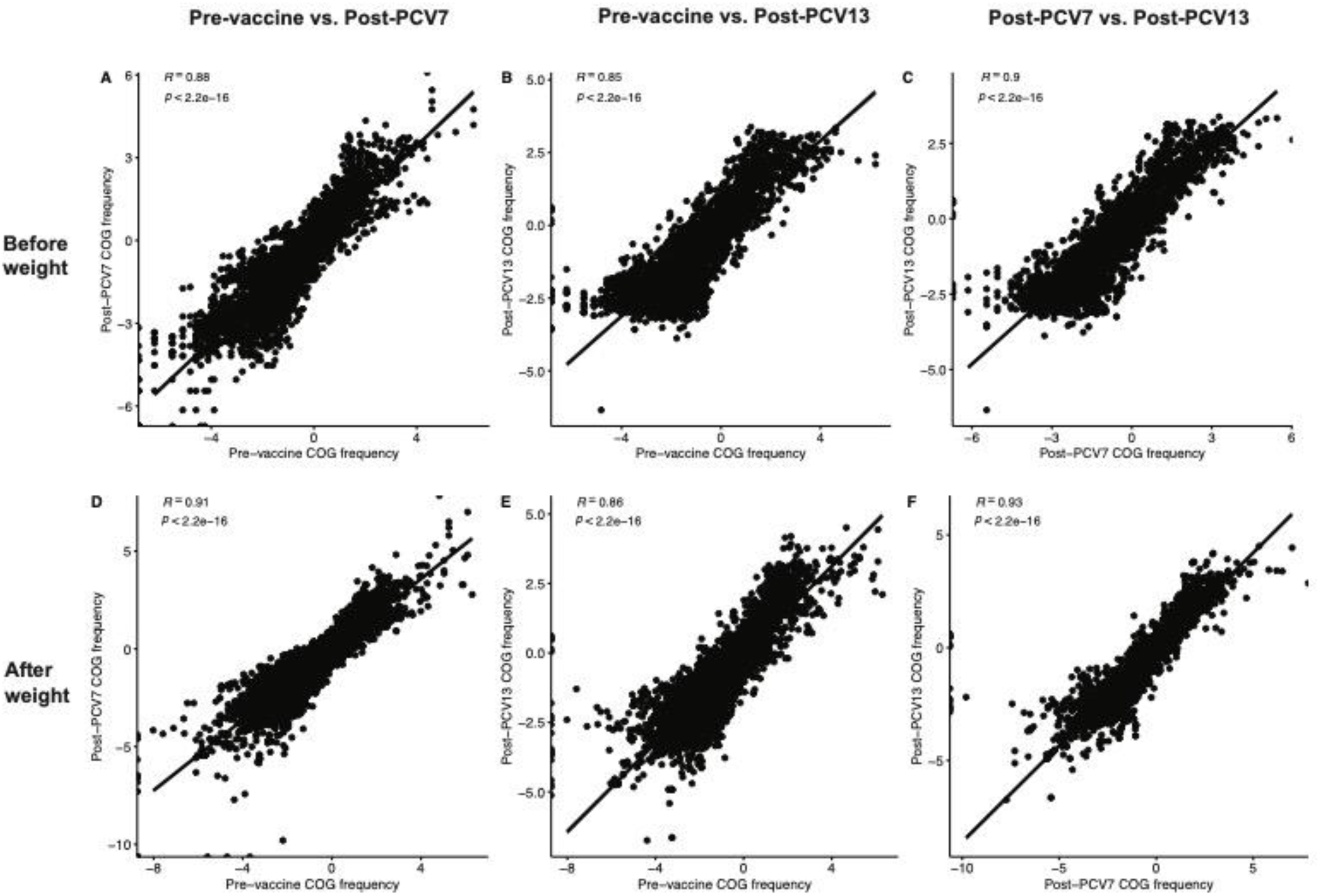
Correlation of logit-transformed accessory gene frequency between different epochs in invasive data before and after weights. A, B, C show the correlations before weights in the IPD data for pre-vaccine vs. post-PCV7 (A), pre-vaccine vs. post-PCV13 (B), and post-PCV7 vs. post-PCV13 (C). D, E, F show the correlations after weights in the IPD data for different epochs. We observed that the weight improved the correlations in the IPD data with both visually tighter scatterplots and improved correlations after weights. COG, accessory clusters of orthologous genes; R, correlation coefficient; PCV7, 7-valent pneumococcal conjugate vaccine; PCV13, 13-valent pneumococcal conjugate vaccine.

**Supplemental Figure 3.**
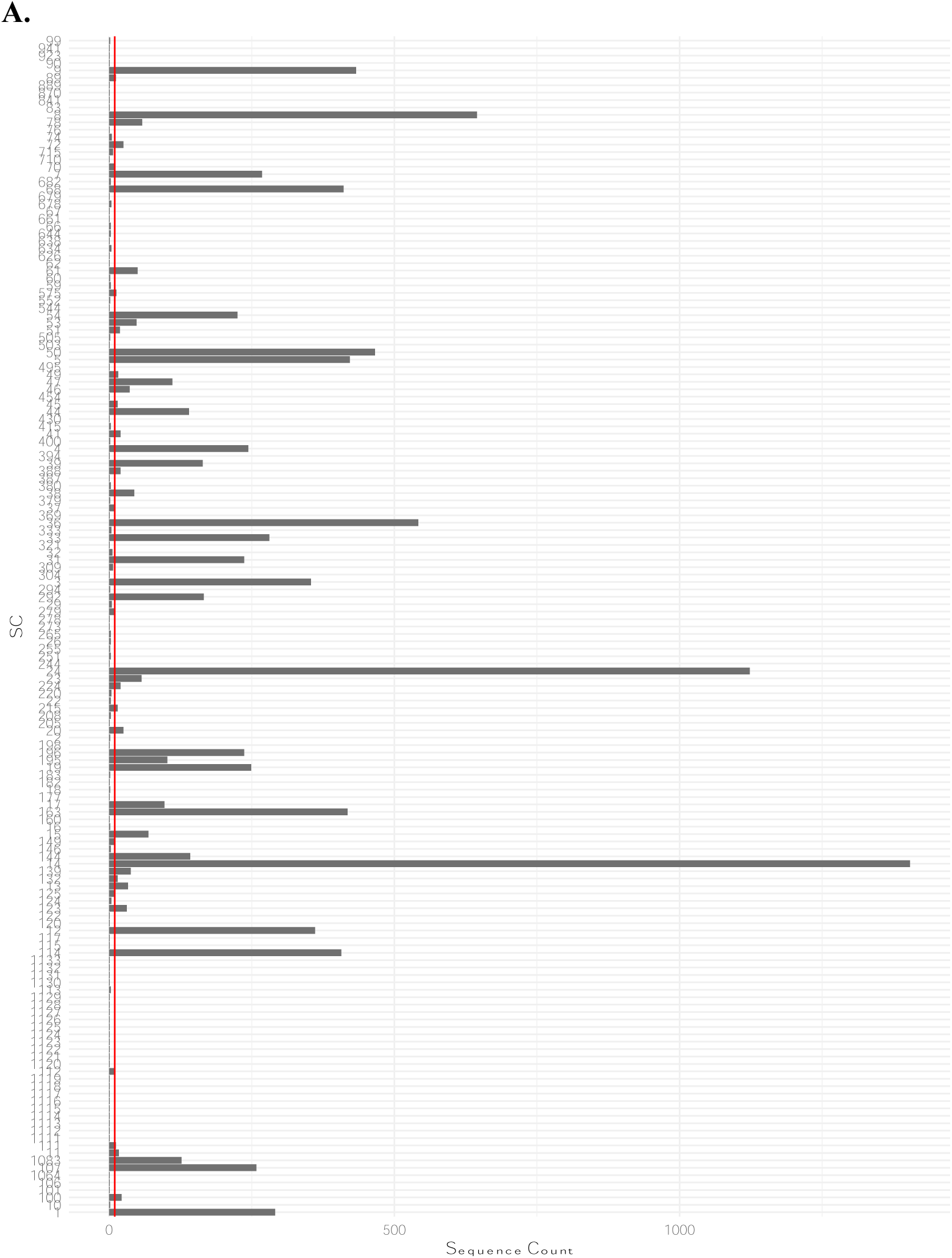
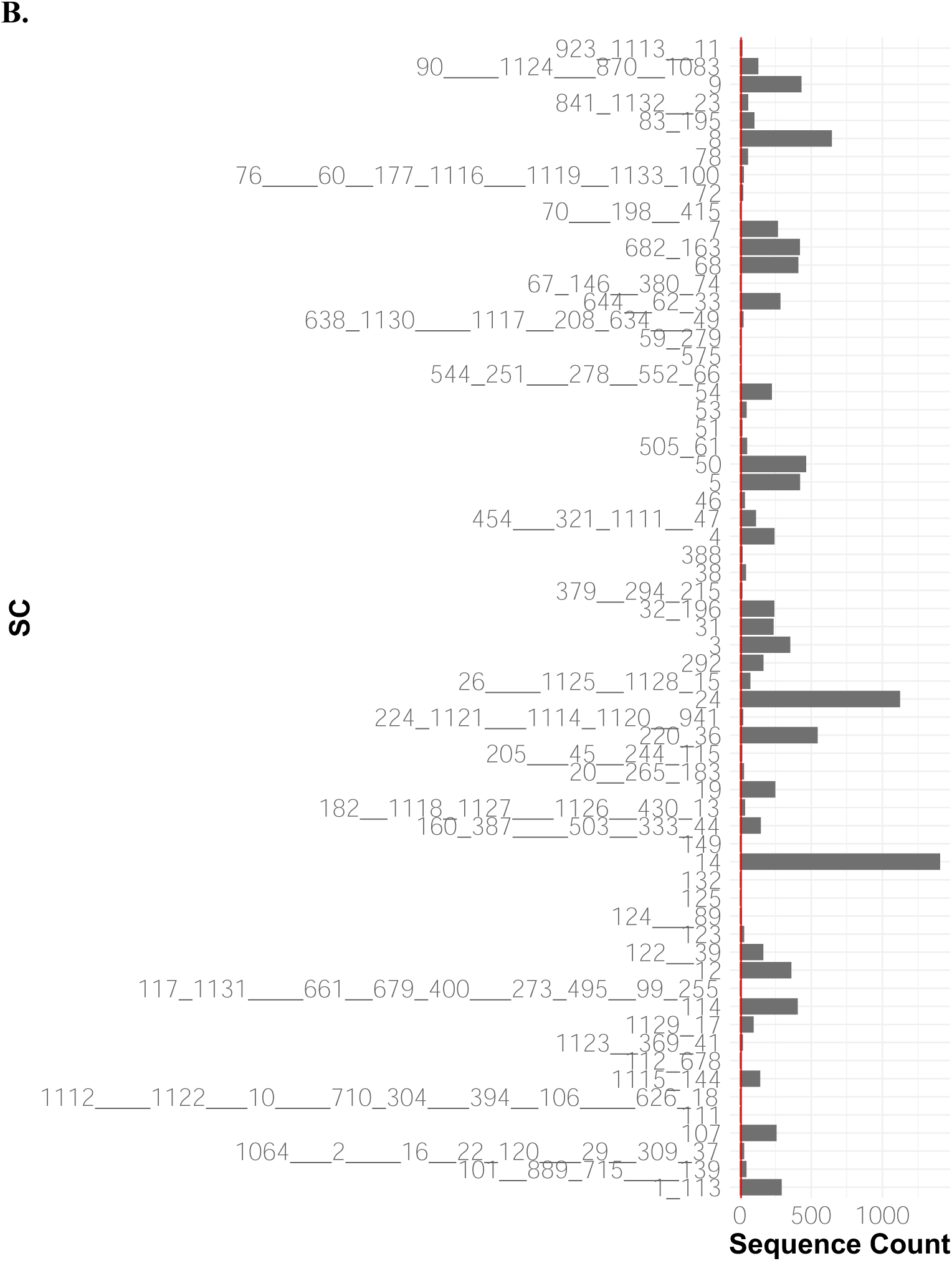
Sequence count in each strain before and after combining small GPSCs. A is the sequence count before the application of weights, and B is after. The name of the strain indicated the original GPSC called by PopPUNK. The underscore connection indicated the combining process by which GPSCs were combined together with minimal accessory frequency distance. The number of underscores indicated at which round out of the 4 rounds the two GPSCs were combined, for example, the connection by one underscore sign indicated that the two GPSCs were combined at the first round. The vertical redline indicates a count of 10 sequences. SC, sequence cluster.

**Supplemental Figure 4.**
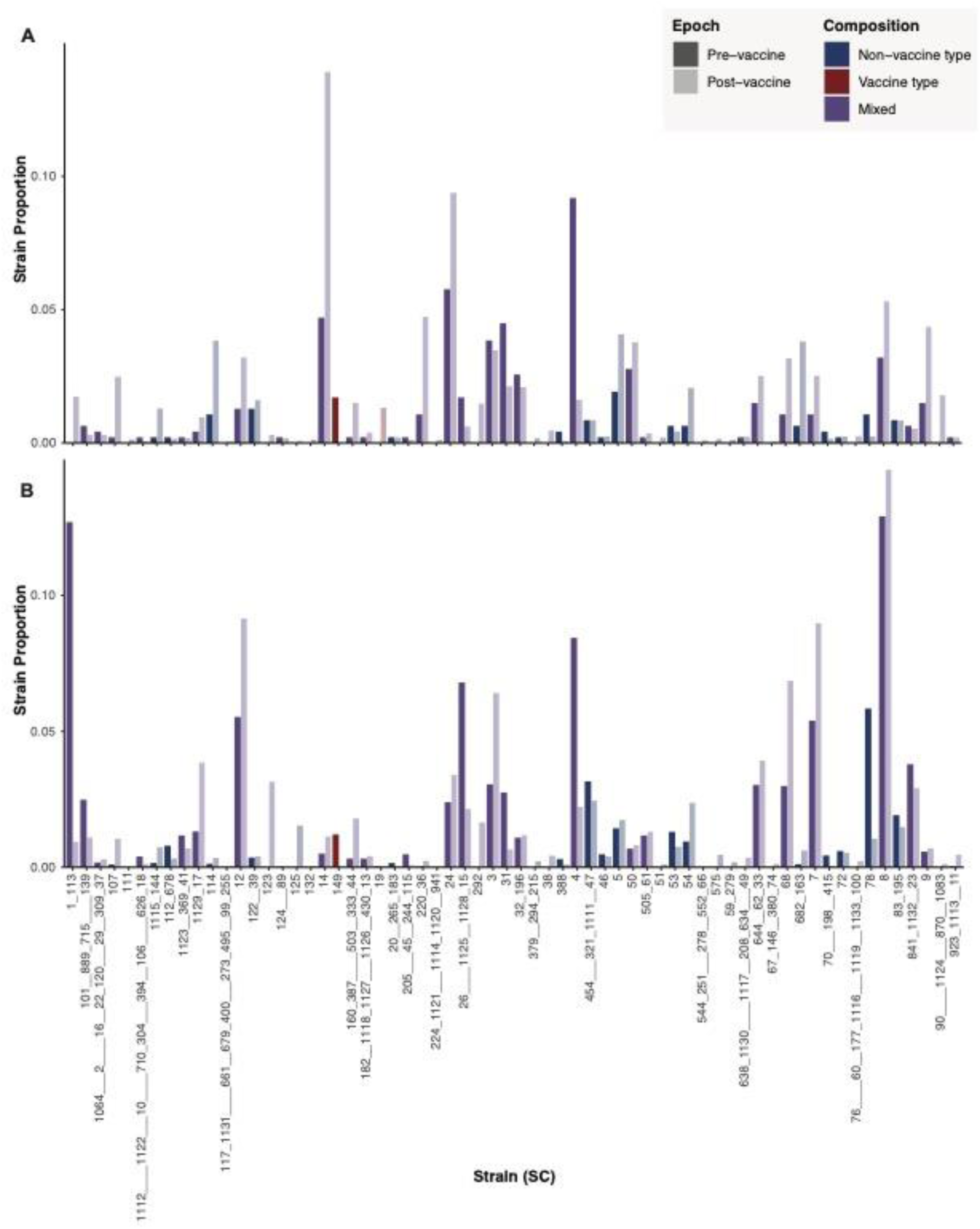
Strain proportion before and after weights considering serotype 3 as NVT. A is the strain proportion before the application of weights, and B is after. The name of the strain indicated the original GPSC called by PopPUNK. The underscore connection indicated the combining process by which GPSCs were combined together with minimal accessory frequency distance. The number of underscores indicated at which round out of the 4 rounds the two GPSCs were combined, for example, the connection by one underscore sign indicated that the two GPSCs were combined at the first round. The weights generally reduced the vaccine types in the strain and increased non-vaccine types. The mixed strains could be increased or decreased depending on the proportion of vaccine/non-vaccine serotypes in a mixed strain.

**Supplemental Figure 5.**
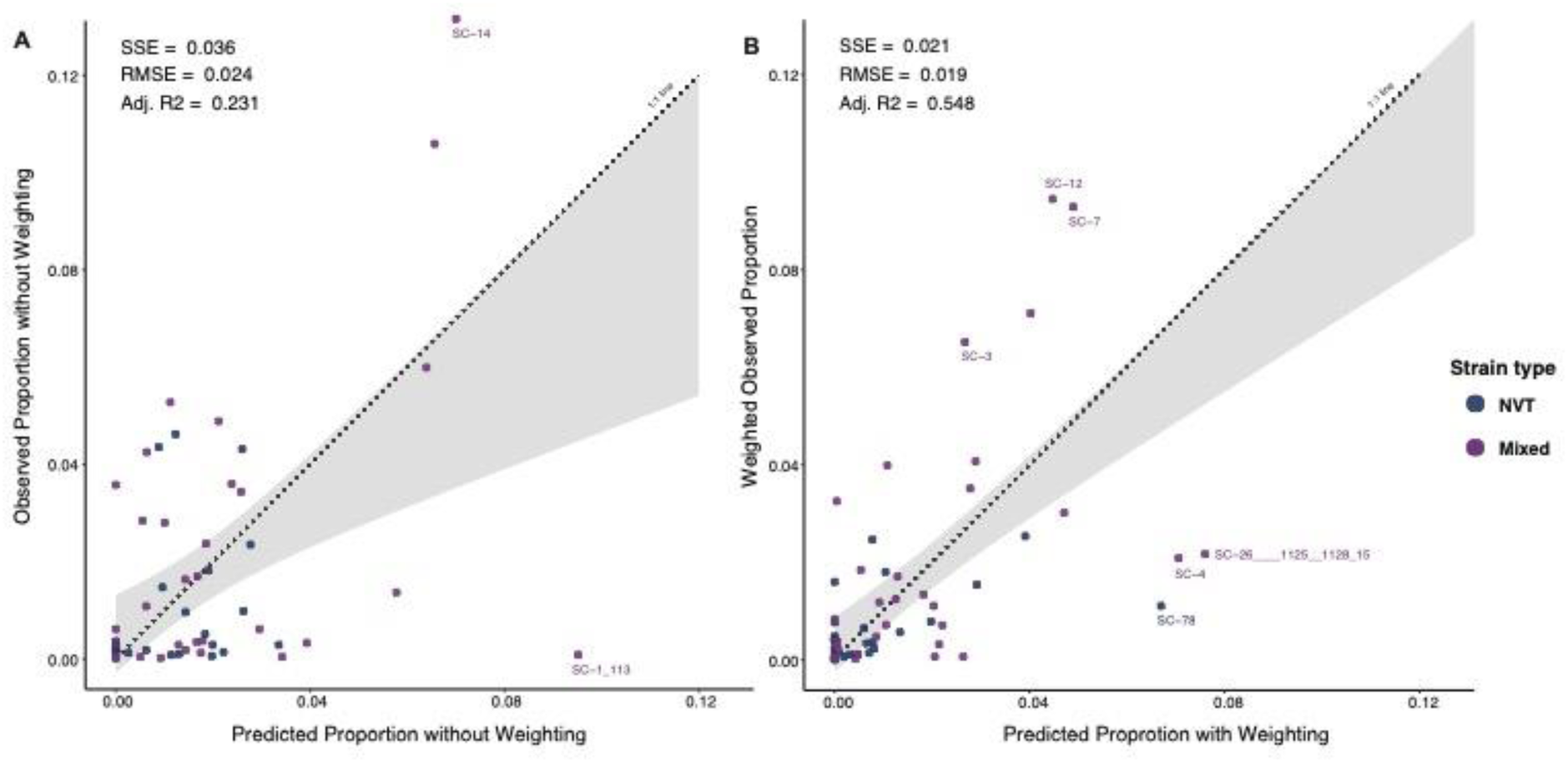
Sensitivity analysis on NFDS prediction model when treating serotype 3 as an NVT. Scatterplot of observed versus predicted proportion of 58 strains at post-PCV13 equilibrium is based on quadratic programming. These 58 strains contained at least 1 NVT isolate pre-vaccine or were imputed with one isolate pre-vaccine. Perfect predictions fall on the dotted line of equality (1:1 line). The shaded gray region shows the confidence interval from the linear regression model used to test for deviation of the observed versus predicted values compared with the 1:1 line. A is the prediction based on the IPD data before the application of weights, and B is after weights. Outliers are defined as the difference between their predicted and observed proportions that is >1.5 times the interquartile range of the distribution of predicted and observed proportion differences. Two outliers were reported before weights applied, including SC-14 (99.9% of serotype 3) and SC-1_113 (92.8% of serotype 19A and 5% of 19F). After weights, the outliers with higher observed proportion than the predicted are SC-3, SC-12 and SC-7, of which the SC-3 contained very diverse serotypes, the SC-12 contained 93.9% of serotype 15A, and the SC-7 contained 38.8% of 23A and 60.1% of 23B. The outliers with higher predicted proportion than the observed included SC-4 (60.2% of 15BC and 37.7% of 19A), SC-26___1125__1128_15 (78.4% of 6C), and SC-78 (94.8% of 6C). NVT, non-vaccine type; Mixed, strain containing both vaccine type and non-vaccine type; SC, sequence cluster or strain; SSE, sum of squares due to error; RMSE, root mean square error; Adj. R2, adjusted R-squared; NFDS, negative frequency-dependent selection; PCV13, 13-valent pneumococcal conjugate vaccine.

